# ColocQuiaL: A QTL-GWAS colocalization pipeline

**DOI:** 10.1101/2021.11.05.21265991

**Authors:** Brian Y. Chen, William P. Bone, Kimberly Lorenz, Michael Levin, Marylyn D. Ritchie, Benjamin F. Voight

**Affiliations:** School of Arts and Sciences, University of Pennsylvania, Philadelphia, PA, USA; Genomics and Computational Biology Graduate Group, Perelman School of Medicine, University of Pennsylvania, Philadelphia, PA, USA; Department of Systems Pharmacology and Translational Therapeutics, Perelman School of Medicine, University of Pennsylvania, Philadelphia, PA, USA; Department of Genetics, University of Pennsylvania, Philadelphia, PA, USA; Corporal Michael J. Crescenz VA Medical Center, Philadelphia, PA, USA; Department of Surgery, Perelman School of Medicine, University of Pennsylvania, Philadelphia, PA, USA; Division of Cardiovascular Medicine, Department of Medicine, University of Pennsylvania Perelman School of Medicine, Philadelphia, PA, USA; Institute for Biomedical Informatics, Perelman School of Medicine, University of Pennsylvania, Philadelphia, PA, USA; Center for Precision Medicine, Perelman School of Medicine, University of Pennsylvania, Philadelphia, PA, USA; Institute for Translational Medicine and Therapeutics, Perelman School of Medicine, University of Pennsylvania, Philadelphia, PA, USA

## Abstract

**Summary:** Identifying genomic features responsible for genome-wide association study (GWAS) signals has proven to be a difficult challenge; many researchers have turned to colocalization analysis of GWAS signals with expression quantitative trait loci (eQTL) and splicing quantitative trait loci (sQTL) to connect GWAS signals to candidate causal genes. The ColocQuiaL pipeline provides a framework to perform these colocalization analyses at scale across the genome and returns summary files and locus visualization plots to allow for detailed review of the results. As an example, we used ColocQuiaL to perform colocalization between the latest type 2 diabetes GWAS data and Genotype-Tissue Expression (GTEx) v8 single-tissue eQTL and sQTL data.

**Availability and Implementation:** ColocQuiaL is primarily written in R and is freely available at github: https://github.com/bychen9/eQTL_colocalizer.

## Introduction

Genome-wide association studies (GWAS) conducted on large populations have identified a plethora of associations between genetic variation and complex traits/diseases in humans^1^. From this collection of predominantly non-coding variants, a central challenge has become identifying which genomic features at each locus ultimately influence the phenotype of interest. This insight is a key barrier to initiate functional follow-up experiments.

One source of data that can be used to link GWAS associations to a predicted gene of action is by connecting them with molecular phenotype quantitative trait loci (QTLs). A well-powered source of two important types of QTLs – those associated with variation in expression of transcripts (eQTLs) and proportion of alternatively spliced transcripts (sQTLs) – was reported across >40 tissues by the Genotype-Tissue Expression (GTEx) project^2^. To connect trait signals to these data and identify potential candidate genes, the community has turned to perform statistical colocalization – an approach designed to infer if the association signals between a complex trait and QTL are tagged by the same genetic variant(s)^3^.

To provide a common, reproducible framework to perform colocalization analyses between QTL and complex trait data at moderate computational scale, we present here an implementation, ColocQuiaL, which allows for the rapid execution of colocalization analysis for GWAS signals from a summary statistics file with all GTEx QTL signals. As a proof of concept, we applied it to the largest catalog of lead associations and summary data for type 2 diabetes (T2D) and the GTEx v8 single-tissue eQTLs or sQTLs datasets^4,5^.

## ColocQuiaL

The motivation underlying the development of ColocQuiaL was the need to perform and visualize the results from a large number (10,000+) of colocalization analyses between signals for one (or more) complex traits and the catalog of available QTL data in GTEx (**Fig. 1**). As such, ColocQuiaL automates the execution of COLOC to perform colocalization analyses between GWAS signals for any trait of interest and GTEx single-tissue eQTL and sQTL signals^3^. The input to ColocQuiaL can be a single locus, a list of loci of interest, or across the entire genome (**Fig. 1**). Users can specify the lead SNPs and the genomic intervals of the colocalization analysis based on prior knowledge of the loci, or they can perform more general analyses by supplying the GWAS summary statistics file and their preferred definition of significant P-values and independent loci via an interface with PLINK^6^. In all these scenarios, ColocQuiaL will perform a colocalization analysis between each single-tissue eQTL or sQTL signal for which the lead SNP is a significant QTL and the GWAS signal at the locus.

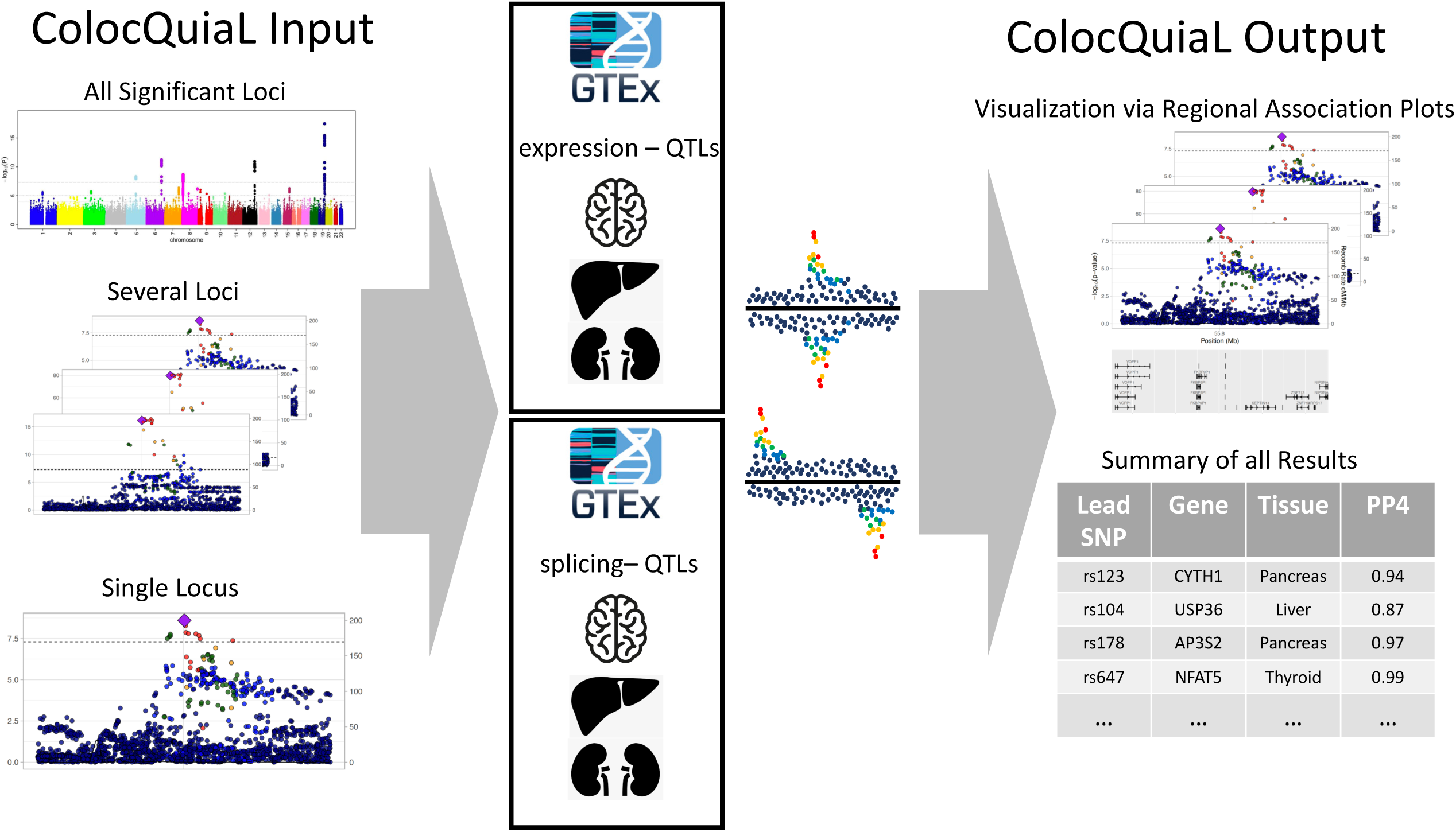

ColocQuiaL generates output files to allow for both manual review of individual colocalization analyses and quick review of all the analyses performed (**Fig. 1**). The majority of these output files are deposited in the lead SNP specific directories. The COLOC results and intermediary files for each colocalization analysis at the lead SNP will all be saved to these directories. These directories will also include regional association plots for each QTL-tissue signal involved in a colocalization analysis and the GWAS trait signal at the locus. Finally, ColocQuiaL generates a summary output file that contains all of the locus level posterior probabilities for the COLOC analyses of the ColocQuiaL run.

The ColocQuiaL pipeline is written in R (v3.6.3 or later) and bash. We implemented a version of ColocQuiaL that is parallelized at the lead SNP level via the LSF workload submission system and an in-series version that can be modified for other job submission systems. ColocQuiaL also interfaces with the following standard bioinformatic tools PLINK (v 1.90Beta45), bedtools (v2.29.1) and Tabix (0.2.5)^6–8^. In order to run the pipeline, user will need to download the publicly available GTEx v8 single-tissue files from the GTEx Portal, and configure a small number of dependency files. Detailed instructions on how to download these data, and configure the dependency files are all available at (https://github.com/bychen9/eQTL_colocalizer).

## Usage Scenario

As a use case, we used ColocQuiaL to perform colocalization analysis of all reported independent T2D genome-wide signals reported recently in Mahajan et al. 2020 with GTEx single-tissue eQTLs and sQTLs using the Vujkovic et. al 2020 T2D summary statistics^4,5^. Across the 520 T2D lead SNPs we found 278 colocalized (PP4/(PP3+PP4)>=0.8) with one or more eQTL signals and 148 colocalized with one or more sQTLs. These colocalizing signals represent 766 genes and 47 tissues among the eQTLs and 268 genes and 48 tissues among the sQTLs.

In total, we performed 9,563 colocalizations between T2D signals and eQTL signals and 38,994 between T2D signals and sQTL signals. We performed this on a PowerEdge R630 Server (2.2Ghz Xeon E5-2699 v4 Dual 22-Core, 512Gb memory) using the lead SNP parallelized version of ColocQuiaL. The median run time and median maximum memory usage for each lead SNP job was 10 minutes 1 seconds and 17.66 GBs for the eQTLs and 7 minutes 49 seconds and 16.56 GBs for sQTLs. Both eQTLs and sQTLs had a small number of outlier lead SNPs that were significant for a much larger number of eQTLs/sQTLs signals in GTEx than the average lead SNP, with the maximum number of colocalizations required for an eQTL lead SNP being 343 and 2,561 for an sQTL lead SNP.

Our results show these T2D GWAS signals colocalize with QTL signals for many of the genes one would expect and replicate recent T2D colocalization studies. We found three maturity-onset diabetes of the young (MODY) gene QTLs colocalized with T2D signals. One MODY gene, *KCNJ11*, had both an eQTL and an sQTL signal that colocalized with T2D signals^9^. We also compared our findings to a predicted causal genes list for T2D and found that T2D signals colocalized with eQTL or sQTL signals for 22 out of the 58 genes^10^. Finally, we compared our results to the recently published T2D QTL colocalization result from Gloudemans et al. 2021 – colocalization of T2D and insulin resistance GWAS data with eQTLs and sQTLs from a subset of GTEx tissues – and Alonso et al. 2021 – colocalization of T2D GWAS data with islets of Langerhans eQTLs^11,12^. We found that our results replicate 24 of 46 genes from Gloudemans et al. 2021 including *PLEKHA1, AP3S2, HMG20A* and 16 of the 31 genes from Alonso et al. 2021 including *HMBS, PCBD1*, and *USP36*.

## Supporting information

T2D_GTEx_eQTL_condPP4_0.8

T2D_GTEx_sQTL_condPP4_0.8

Replicated_T2D_colocalizations

## Data Availability

All data produced in the present work are contained in the manuscript and supplemental materials

